# Knowledge, Attitude, and Practices towards COVID-19 Infection and Prevention Measures among Medical Students

**DOI:** 10.1101/2024.02.12.24302741

**Authors:** Jade Monica Marie J. Ruyeras, Priya Kaur V. Basi, Louise Anne C. Cañete, Neal Abram M. Capoy, Mary Ysabelle S. Castillo, Cristine Jayne T. Colonia, Bea Lou Marie E. Gantuangco, Primo Andrio V. Jumamil, Hubert Paul S. Mantilla, Giovanni Sergius C. Talili, Riana Camille G. Untal

## Abstract

The COVID-19 pandemic has rapidly led to an unprecedented health threat worldwide. During this time, disease prevention is considered to be the best way for general health protection. This is achieved through public health education by extending proper knowledge, promoting an optimistic attitude, and keeping the public compliant with preventive measures. As components of the healthcare system, medical students should also play a role in disease prevention more so in the Philippines where the Department of Health has called upon medical graduates to render services in response to the national emergency. In this study, the knowledge, attitude, and practices (KAP) of medical students from Cebu Institute of Medicine (CIM) towards COVID-19 infection and preventive measures were assessed. Demographic factors, their respective effect size on KAP, as well as the relationship between KAP variables were determined. The revised questionnaire, drafted based on qualitative and quantitative validity tests, was then used for the pilot study to generate the final questionnaire. Responses from participants underwent descriptive and correlational analysis. The results showed that the majority of the medical students of CIM have adequate knowledge (78.24%), positive attitude (80.68%), and good practices (94.38%) towards COVID-19 infection and prevention measures. Females have a significant association (p-value = 0.03) with better practices than male counterparts. Knowledge (p-value = 0.004) and attitude (p-value = 0.003) also showed significant correlation with practices, implying that knowledge and attitude play a role in shaping compliance to health practices. Therefore, health interventions should aim to disseminate accurate, evidence-based information and improve attitude towards the implemented precautionary measures in order to increase effectiveness of policies.

## INTRODUCTION

The SARS-CoV-2 is a novel coronavirus that causes coronavirus 2019 or COVID-19, a highly communicable and pathogenic viral infection that produces mild to moderate respiratory illness [1,2]. COVID-19 emerged from the city of Wuhan in China and became a worldwide problem in the year 2020 [2]. In the Philippines, the first COVID-19 case was confirmed by the Department of Health (DOH) on January 30, 2020. The number of cases rose exponentially as the months went by, reaching 491,258 confirmed cases as of January 12, 2021 [3,4]. Based on the DOH national assessment, the Philippines remains in Stage 2, localized community transmission with several localities exhibiting higher transmission intensity [4].

Due to the novelty of the virus, lack of evidence-based cure, and limited vaccine roll-out in our locality, preventive measures have become increasingly important for general health protection. In the Philippine context, our government enforced preventive measures such as community quarantine protocols, especially in regions with a rising number of cases in order to contain further transmission. These protocols include home quarantine, lockdowns, suspension of public transportation systems, and restricting travel to other places [3]. Personal safety measures, such as social distancing, wearing of face masks, proper hand hygiene and respiratory etiquette were implemented as well [5].

Despite the toll the viral infection has taken on the whole world, some individuals continue to underestimate the risk of transmission and remain poorly compliant to health protocols [6]. Effective promotion of precautionary measures would require the identification of factors associated with the practices [7]. In a study by Webster et al. (2020), possible determinants for adherence to quarantine protocols during previous infectious disease outbreaks include demographic characteristics, knowledge on the disease and quarantine protocol, sociocultural factors, perceptions on disease risk and quarantine benefit, health center functionality, length of quarantine and trust in government response [8]. Assessment of the knowledge, attitude, and practices (KAP) towards COVID-19 infection and prevention aids in the identification of misconceptions and construction of possible health intervention strategies that influence KAP. This information would be important to healthcare professionals, service providers and medical students who are at risk for contact with infected patients and who also play a role in educating the public with accurate information and increasing their awareness of COVID-19. Prior KAP studies among medical students show the relationship between these variables wherein a higher knowledge level and a positive attitude would correlate with better practice of preventive measures [9, 10]. However, a KAP study among medical students has not yet been conducted in Cebu City which was considered to be one the COVID-19 hotspots in the Philippines [11].

In this study, the knowledge level, attitude, and practices towards COVID-19 infection and prevention measures among medical students in Cebu Institute of Medicine are assessed. KAP would also be analyzed with respect to the demographic profile of respondents in order to identify possible sociodemographic factors and determine their effect size on KAP. The relationship between the KAP variables with regards to COVID-19 infection and prevention would also be determined in this study.

## METHODOLOGY

### Study Design and Setting

A cross-sectional survey was employed to assess the knowledge, attitude, and practices towards COVID-19 infection and prevention measures of Cebu Institute of Medicine students. These factors which may affect compliance to the recommended measures that prevent transmission were assessed through a researcher-generated questionnaire. Data was collected through online dissemination of the questionnaire through Google Forms with consequent descriptive and correlational analysis.

### Study Population

The study population are bonafide medical students of Cebu Institute of Medicine who were enrolled for the second semester of school year 2020-2021. Medical students of all year levels from first to fourth year were covered in this study. As of the second semester of school year 2020-2021, there are 608 enrolled students in CIM: 111 in the first year level, 173 in the second year level, 168 in the third year level, and 156 in the fourth year level.

### Inclusion Criteria and Exclusion Criteria

The inclusion criteria were being a bonafide student of CIM enrolled for school year 2020-2021, having internet access, and voluntary participation.

### Sample Size Calculation

The sample size was calculated from a known population with the Slovine’s formula:

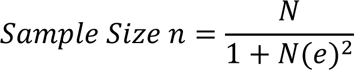

Where the population size (N) is 111 first year students, 173 second year students, 168 third year students, and 156 fourth year students. Using a margin of error (e) of 0.05 or 5% with 95% confidence level, the calculated minimum required sample size (n) should be 44 first year students, 69 second year students, 67 third year students, and 62 fourth year students as shown in Table 1.

**TABLE 1.**
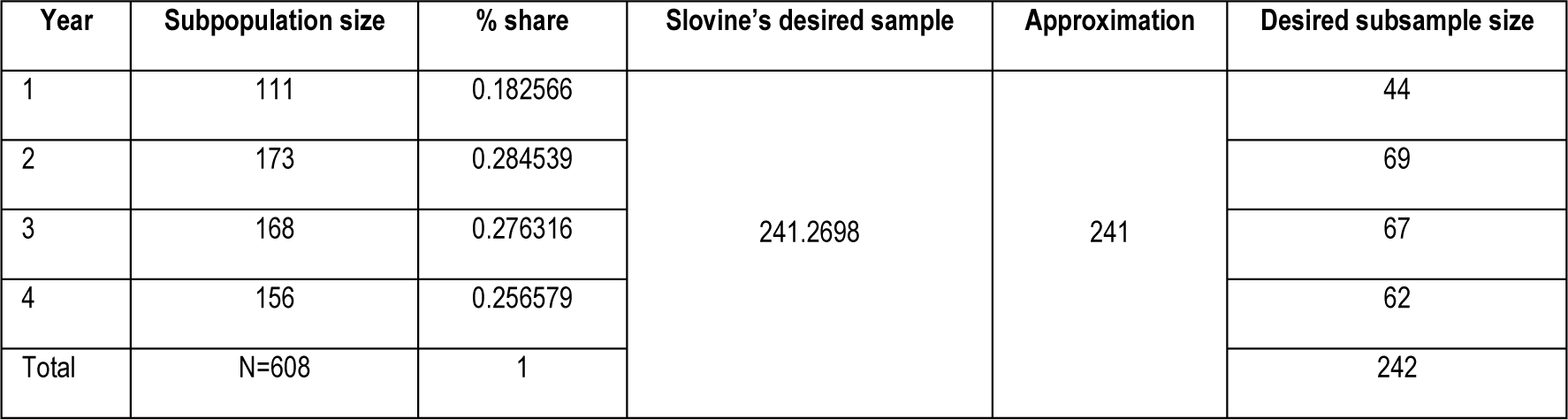
DESIRED SUBSAMPLE SIZE.

| Year | Subpopulation size | % share | Slovine's desired sample | Approximation | Desired subsample size |
| --- | --- | --- | --- | --- | --- |
| 1 | 111 | 0.182566 | 241.2698 | 241 | 44 |
| 2 | 173 | 0.284539 |  |  | 69 |
| 3 | 168 | 0.276316 |  |  | 67 |
| 4 | 156 | 0.256579 |  |  | 62 |
| Total | N=608 | 1 |  |  | 242 |

### Data Collection Process

The participants of the study were contacted through their emails and social media. Class representatives for each year level were tapped for distribution of the questionnaire link to their respective batches. They were briefed about the nature of the study and asked for their consent to participate. An online questionnaire, via Google Forms, was then administered to them. Their responses were immediately accounted for once they have submitted the form.

### Data Collection Tool

The questionnaire was adapted and modified from previously published papers regarding the knowledge, attitude, and practices of the public towards outbreaks especially COVID-19. Furthermore, an 80% cut-off value was applied for all the questionnaire sections based on these aforementioned literature [12, 13, 14, 15]. The questionnaire was divided into four parts: demographic characteristics, knowledge, attitude, and practices. The demographics section consists of 5 items which included sex, year level, hometown, quarantine location, and sources of information regarding COVID-19.

The knowledge section consists of 8 multiple-choice items which included etiologic agent, incubation period, mode of transmission, presenting symptoms, high-risk population for severe outcome, preventive measures, possible complications, and mortality rate. A point is given for each correct answer and no points are given for every incorrect answer such that the score for this portion would range from 0 to 8. Adequate knowledge is defined by a score of at least 6 while inadequate knowledge is defined by a score below 6.

The attitude section consists of 6 items that assessed belief in a personal responsibility to take safety measures to prevent COVID-19 transmission, belief of virus transmission from an asymptomatic patient, belief in hand washing and wearing of masks as a form of infection prevention, belief in the protective benefits of COVID-19 vaccine, belief that the COVID-19 vaccine would be accessible to most people, and belief that COVID-19 would be completely controlled. These items were evaluated using a 5-point Likert scale. The options for each question were strongly agree, agree, neutral, disagree, and strongly disagree which are given 5, 4, 3, 2, and 1 point respectively. The total score for this portion would then range from 6 to 30. Positive attitude is defined by a score of at least 24 while negative attitude is defined by a score below 24.

The practices section consists of 12 items that assessed the frequency of performing COVID-19 precautionary measures including wearing of face mask, wearing of face shield, washing hands, using disinfectants, avoiding public gatherings, staying at home as much as possible, avoiding eating outside in crowded places, closely monitoring personal physical health, closely monitoring the physical health of people around you, persuading people around you to follow precautionary guidelines, following social distancing protocols, and self-isolating when feeling unwell or manifesting COVID-19 symptoms. These items were evaluated using a 5-point Likert scale. The options for each question are always, often, sometimes, rarely, and never which were given 5, 4, 3, 2, and 1 point respectively. The total score for this portion would then range from 12 to 60. Good practice is defined by a score of at least 48 while bad practice is defined by a score below 48.

### Validity and Reliability of the Data Collection Tool

The initial draft of the survey questionnaire was submitted to 6 academic experts who are knowledgeable in the area. In the qualitative content validity method, experts gave their feedback on grammar, use of appropriate and correct words, application of correct and proper order of words in items, and appropriate scoring. In the quantitative content validity method, each item was rated as not necessary, useful but not essential, or essential. Validity was then assessed by determining the content validity ratio (CVR) which was calculated through the given formula:

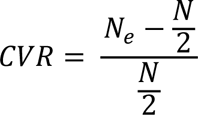

Where Ne is the number of panelists who indicated that an item is “essential” and N is the total number of panelists. The minimum value of CVR for item inclusion was determined to be at least 0.78 [16].

Considering the recommendations and consensus of all experts, the final questionnaire was formulated and utilized for pilot testing in 35 individuals to confirm the reliability of the questionnaire. Data from the pilot study was subjected to reliability coefficient analysis to evaluate internal consistency which can be estimated using the coefficient alpha or Cronbach’s alpha which is calculated through the given formula:

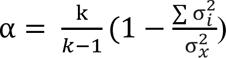

Where k is the number of items, σ_*i*_^2^ is the variance of item i, and σ_*x*_^2^ is the total variance of questionnaire x. Cronbach’s alpha ranges from 0 which indicates no internal consistency to 1 which indicates perfect internal consistency [17]. Adequate internal consistency is demarcated by a Cronbach’s alpha value of at least 0.70 [18].

### Data Analysis

Data generated from the online survey through Google Forms was downloaded as an Excel file. The data was then imported to Statistical Package for Social Sciences (SPSS) for analysis.

### Statistical Treatment

Descriptive statistical analysis was performed for the demographic characteristics of the respondents which was presented through frequency and simple percentage. A chi-square test for independence will be used to measure the association between the demographics with knowledge, attitude, and practice variables. To identify the effect size of the factors significantly associated with adequate knowledge, positive attitude and appropriate practice, a binary logistic regression was performed and the odds ratio was also reported. Pearson rho correlation was also used for measuring the intra variable relationships between knowledge, attitude and practices. All hypotheses testing was performed using the level of significance of 0.05.

### Level of Significance

The significance level α was set to 0.05. Thus, for any given hypothesis test, a p-value less than 0.05 is deemed to be statistically significant. For the Pearson correlation analysis, significant correlation is shown at the level of p = 0.01.

### Ethical Consideration

A full consent to participate was obtained from the respondents prior to study. The respondents were briefed regarding the nature and purpose of the study with provision of sufficient information to fully understand the implications of participation. The respondents were given absolute freedom to decline the recruitment. The confidentiality of the given information and anonymity of respondents is of utmost importance, ensuring the protection of their privacy.

#### Informed Consent

Before the respondents answered the online questionnaire, consent was asked from them. The informed consent form contained the title of the study, the proponents, and the goals of the study. Participation of the respondents was completely voluntary. They may opt not to join the survey with no negative consequences. By completing and submitting the form, participants indicated that they have read the consent and agreed to the terms as described.

#### Confidentiality and Security of Information

Any information provided by the respondents will be kept confidential and will be used only for the purposes of completing this study. Information and responses of the respondents were processed and did not include the name or any other individual information by which any respondent could be identified. All responses and records will be kept secure by the principal investigators of the study.

#### Benefits

This study did not require mandatory participation of the Cebu Institute of Medicine students. The participants had the right to decline or to withdraw from the study at any time if they wished to do so. They also the right to know the results of this study.

#### Protection

Any personal information gathered will not be released without the respondent’s consent. The researchers are willing to address any concerns that would possibly harm the respondents by participating in this study.

## RESULTS

A total of 409 Cebu Institute of Medicine students participated in the survey to determine the knowledge levels, attitude, and practices towards COVID-19 infection and prevention measures among medical students. The results were tabulated and analyzed. Likewise, the sociodemographic profiles and its association with the knowledge levels, attitude, and practices were also presented.

### Demographic Characteristics

Table 2 shows the demographic characteristics of the respondents which showed that among the 409 respondents, majority were females (64.55%). Most of the participants were from the 2^nd^ year level (29.34%) while 1^st^ year, 3^rd^ year, and 4^th^ year medical students comprised 19.32%, 24.94%, and 26.41% respectively. About half (50.37%) were residing within Cebu City during the entire quarantine period from March 2020 up to the time of the survey (50.37%) but most of the respondents came from hometowns outside of Cebu City (63.08%).

**TABLE 2.** DEMOGRAPHIC CHARACTERISTICS OF THE CIM STUDENT RESPONDENTS.

| Profile | Frequency (f)<br>n=409 | Percentage (%) |
| --- | --- | --- |
| **Sex** |  |  |
| Male | 145 | 35.45 |
| Female | 264 | 64.55 |
| **Year Level** |  |  |
| 1st | 79 | 19.32 |
| 2nd | 120 | 29.34 |
| 3rd | 102 | 24.94 |
| 4th | 108 | 26.41 |
| **Within Cebu City during Quarantine** |  |  |
| Yes | 206 | 50.37 |
| No | 203 | 49.63 |
| **Hometown** |  |  |
| Within Cebu City | 150 | 36.67 |
| Outside | 258 | 63.08 |
| **Sources of Information** |  |  |
| Social media | 398 | 97.31 |
| Google | 366 | 89.49 |
| Medical Search Engine | 254 | 62.10 |
| Family Friends | 326 | 79.71 |
| Official websites (WHO, CDC, etc.) | 373 | 91.20 |
| TV | 253 | 61.86 |
| Healthcare Workers | 349 | 85.33 |
| NGOs | 99 | 24.21 |
| Religious Leader | 30 | 7.33 |

As to their source of information regarding COVID-19 infection and prevention, social media (97.31%) was their primary source followed by official websites (91.20%), Google search (89.49%) and healthcare workers (85.33%).

### Knowledge, Attitude, and Practices towards COVID-19 Infection and Prevention

Figure 1 shows that a high number of CIM student respondents are knowledgeable on the etiology, incubation period, symptoms, preventive measures, and complications of COVID-19 wherein 95.11%, 93.89%, 97.07%, 99.51%, and 88.75% respectively were correct. On the other hand, it is noted that the number of students who are knowledgeable on the mode of transmission (69.44%), high-risk population (44.99%), and the case fatality rate (44.01%) is lower. For mode of transmission, 23.72% of the respondents thought that COVID-19 can only be transmitted through respiratory droplets. For the high-risk population, 54.77% of the respondents thought that high-risk populations for severe disease included children and pregnant women aside from the elderly and people with underlying medical conditions. For case fatality rate, 57.21% of the respondents thought that the case fatality rate in Southeast Asia was higher than 1.40. Overall, it can be validated that the majority of the respondents were classified to have adequate knowledge (78.24%).

**FIGURE 1.**
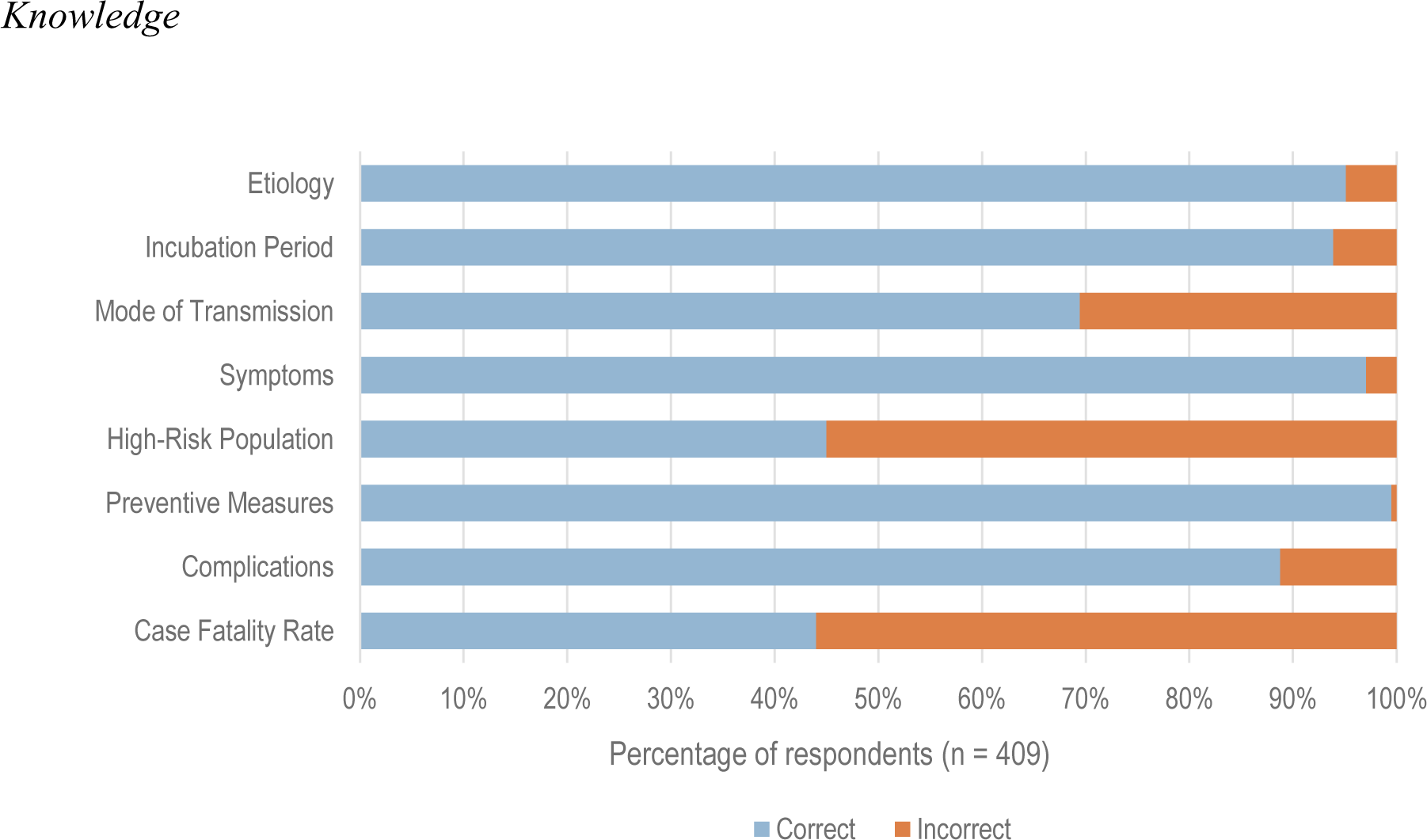
KNOWLEDGE LEVELS ON COVID-19 INFECTION AND PREVENTION OF CIM STUDENTS.

Figure 2 shows that majority of the CIM student respondents have a positive attitude towards COVID-19 infection and prevention as they strongly agree that it is their social responsibility to take safety measures in controlling the spread of infection (91.93%), an asymptomatic patient can transmit the virus to others (88.51%), the COVID-19 vaccine can help them be protected from infection (78%), infection can be prevented through hand washing and wearing of face masks (46.45%), and that the COVID-19 vaccine will be made accessible to most people (32.76%). It should be noted that most of the respondents agreed that COVID-19 will be controlled completely (29.34%) but 28.36% are neutral. Overall, it can be validated that the majority of the respondents were classified to have a positive attitude (80.68%).

**FIGURE 2.**
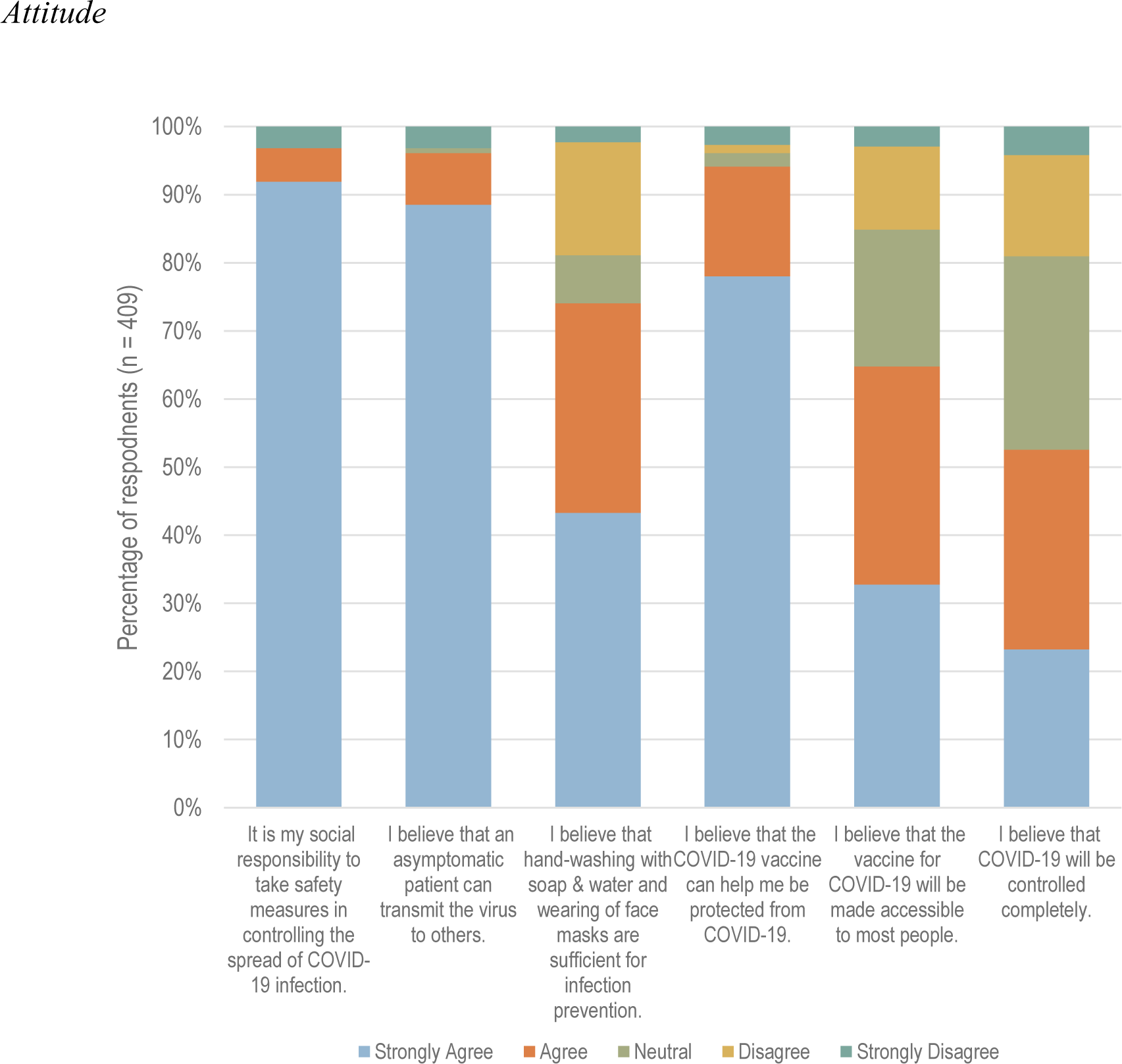
ATTITUDE TOWARDS COVID-19 INFECTION AND PREVENTION OF CIM STUDENTS.

Figure 3 shows that the majority of CIM student respondents always practice precautionary measures against COVID-19 especially wearing of face masks (95.84%), regular washing of hands (81.17%), and using disinfectants (81.17%). Overall, it can be validated that the majority of the respondents were classified to have good practices (94.38%).

**FIGURE 3.**
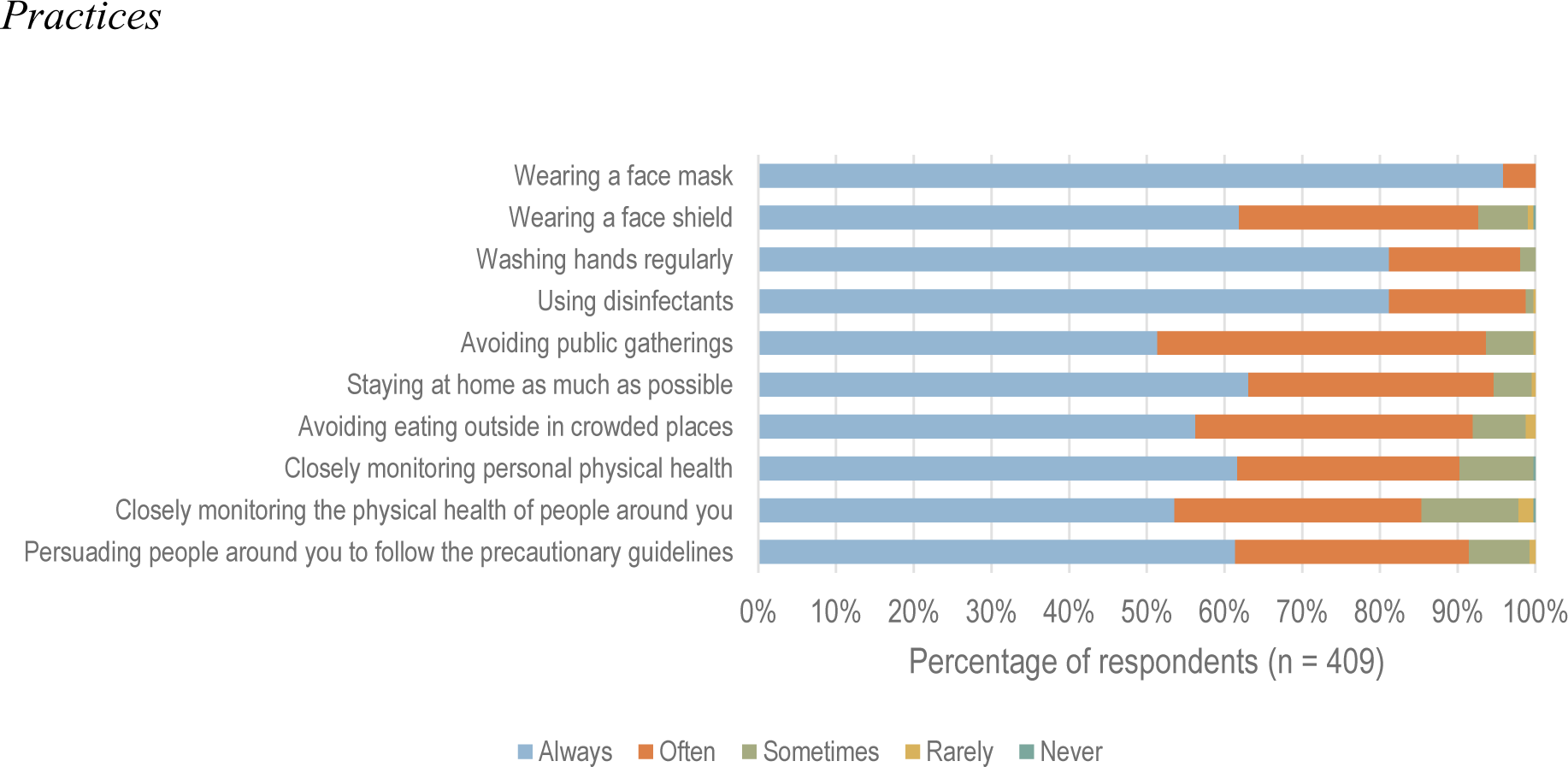
FREQUENCY OF PRACTICING COVID-19 PRECAUTIONARY MEASURES OF CIM STUDENTS.

### Demographic Factors Affecting KAP towards COVID-19 Infection and Prevention

In terms of testing the association between the demographic characteristics of the respondents and their level of KAPs, a Chi-square test was performed. Table 3 succinctly reveals that only sex, referring to being a female or a male, is significantly associated (p-value = 0.03) with the extent of practicing COVID-19 precautionary measures. In fact, there is a significantly higher proportion of females (96.21%) who have better practices than their male counterparts (91.03%).

**TABLE 3:** CHI-SQUARE TEST RESULTS OF THE DEMOGRAPHIC VARIABLES VS KAP.

| Profile | Knowledge |  |  | Attitude |  |  | Practice |  |  |
| --- | --- | --- | --- | --- | --- | --- | --- | --- | --- |
|  | Adequate (320) | Inadequate (89) | p-value | Positive (330) | Negative (79) | p-value | Good (386) | Bad (23) | p-value |
| **Sex** | n (%) | n (%) |  | n (%) | n (%) |  | n (%) | n (%) |  |
| Male | 121 (37.81) | 24 (26.97) | 0.06 | 119 (39.06) | 26 (32.91) | 0.60 | 132 (91.03) | 13 (8.97) | 0.03* |
| Female | 199 (62.19) | 65 (73.03) |  | 211 (63.94) | 53 (67.09) |  | 254 (96.21) | 10 (3.79) |  |
| **Year Level** |  |  |  |  |  |  |  |  |  |
| 1st | 60 (18.75) | 19 (21.35) | 0.78 | 17 (21.52) | 62 (18.79) | 0.08 | 77 (19.95) | 9 (39.13) | 0.32 |
| 2nd | 94 (29.38) | 26 (29.21) |  | 31 (39.24) | 89 (26.97) |  | 114 (29.53) | 6 (26.09) |  |
| 3rd | 78 (24.38) | 78 (24.38) |  | 17 (21.52) | 85 (25.76) |  | 93 (24.09) | 6 (26.09) |  |
| 4th | 88 (27.50) | 20 (22.47) |  | 14 (17.72) | 94 (28.48) |  | 102 (26.42) | 2 (8.70) |  |
| **In Cebu City during Quarantine** |  |  |  |  |  |  |  |  |  |
| Yes | 162 (77.83) | 44 (21.36) | 0.84 | 164 (79.61) | 42 (20.39) | 0.58 | 195 (94.66) | 11 (5.34) | 0.80 |
| No | 158 (78.64) | 45 (22.17) |  | 166 (81.77) | 37 (18.23) |  | 191 (94.09) | 12 (5.91) |  |
| **Hometown** |  |  |  |  |  |  |  |  |  |
| Within Cebu | 117 (36.56) | 33 (37.08) | 0.93 | 118 (35.76) | 32 (40.51) | 0.43 | 143 (95.33) | 7 (4.67) | 0.52 |
| Outside | 203 (63.44) | 56 (62.92) |  | 212 (62.24) | 47 (59.49) |  | 243 (93.82) | 16 (6.18) |  |

\*Association is significant at p-value $\leq 0.05$ level (2-tailed)

Moreover, the rest of the demographic factors did not associate among the KAP measures which suggest that there is no sufficient statistical evidence yet to claim that their impact to the KAPs is statistically material and considerable.

As shown in Table 4, a binary logistic regression was performed to determine the effect size of the different factors on the KAP domains. Being a female student recorded 1.65 times more likely to have adequate knowledge and positive attitude than his male counterpart. Likewise, the 4^th^ year students were 1.35 times more likely to have adequate knowledge while the 2nd year students were 1.74 times more likely to manifest good attitudes than their 3^rd^ year counterparts. Being within the city during the quarantine period portend to promote 1.11 times increased likelihood for students to practice measures against transmission and infection. Students whose permanent residences were located within the city also gave odds of 1.21 to possess a positive attitude more than the odds of those who did not reside in the city. However, all confidence intervals cross 1 (e.g. 95% CI for sex at 0.979, 2.769), which then implies that there is no difference between the groups considered in the study.

**TABLE 4.** BINARY LOGISTIC REGRESSION RESULTS OF THE DEMOGRAPHIC VARIABLES VS KAP.

| Predictor | Knowledge | Attitude | Practice |
| --- | --- | --- | --- |
|  | Adjusted OR (95% CI) | Adjusted OR (95% CI) | Adjusted OR (95% CI) |
| **Sex** |  |  |  |
| Male | reference | Reference | reference |
| Female | 1.647<br>(0.979, 2.769) | 1.150<br>(0.683, 1.934) | 0.40 |
| **Year Level** |  |  |  |
| 1st | 0.972 | 1.371<br>(0.649, 2.896) | 0.268 |
| 2nd | 1.112<br>(0.592, 2.090) | 1.742<br>(0.898, 3.376) | 0.608 |
| 3rd | reference | Reference | Reference |
| 4th | 1.354<br>(0.695, 2.638) | 0.745 | 0.544 |
| **Within Cebu City during Quarantine** |  |  |  |
| Yes | 0.954 | 0.870 | 1.114<br>(0.480 2.585) |
| No | reference | Reference | Reference |
| **Hometown** |  |  |  |
| Within Cebu | 0.991 | 1.207<br>(0.731, 1.995) | 0.735 |
| Outside | reference | Reference | Reference |

OR>1.00 increases the likelihood of an event to occur

### Relationship between Knowledge, Attitude, and Practices

As shown in Table 5, the Pearson-rho correlation was utilized to analyze the relationship between the KAP variables. Knowledge and attitude recorded a positive coefficient of 0.04. However, this relationship was not significant at p-value 0.43 (p > 0.05). This means that with the current statistical evidence, there is no proof to show that knowledge contributed positively to the attitude of the respondents.

**TABLE 5.** CORRELATIONAL ANALYSIS BETWEEN KNOWLEDGE, ATTITUDE, AND PRACTICES TOWARDS COVID-19 INFECTION AND PREVENTION MEASURES AMONG CIM STUDENTS.

| Factors | Correlation | Knowledge | Attitude |
| --- | --- | --- | --- |
| Attitude | Pearson Correlation | 0.039 |  |
|  | Sig. (2-tailed) | 0.428 |  |
|  | N | 409 |  |
| Practice | Pearson Correlation | -0.144 | 0.147 |
|  | Sig. (2-tailed) | 0.004* | 0.003* |
|  | N | 409 | 409 |

\*Correlation is significant at $p\text{-value} \leq 0.01$ level (2-tailed)

However, knowledge and practice showed a significant extent of negative (coefficient =-0.144) relationship as depicted by the p-value of 0.004. This translated that there was sufficient evidence to claim that the increase or decrease of knowledge level contributed to the decline or frequency in practice of the respondents.

Lastly, attitude showed a positive effect to practice as evident from the Pearson coefficient of 0.147 which is also significant (p-value 0.003 < 0.05). This means that there is sufficient evidence to claim that positivity of attitude significantly increased the frequency of practice of the students. This is indicative also that positive attitude translated significantly to better compliance to COVID-19 prevention measures.

## DISCUSSION

The abrupt emergence of the COVID-19 pandemic caused a staggering loss of human life worldwide and led to an unprecedented challenge to all sectors [19]. General health protection is therefore sought after more so during this time of the pandemic. This may be achieved through public health education by extending proper knowledge, promoting an optimistic attitude, and keeping the public compliant with preventive measures [15]. Through the assessment of current public knowledge, attitude, and practices (KAP) towards COVID-19 infection and prevention, it aids in the identification of misconceptions and construction of possible intervention strategies that influence KAP. This information would be important to healthcare professionals, service providers and medical students who are at risk for contact with infected patients and who also play a role in educating the public with accurate information and increasing their awareness of COVID-19.

KAP may be shaped by the source from which our information is derived. In this study, social media was the primary source of information among medical students regarding COVID-19 infection and prevention wherein 97.31% of the respondents utilized it. This is in accordance with similar studies which also identified social media as a major source of information on the influenza and coronavirus pandemics [20, 21]. Due to the increasing use of social media as a means of communication, it has become one of the avenues as to where the general public gathers information regarding COVID-19, its symptomatology, recent researches, and vaccination updates among others [21]. With most of the population staying in their homes due to the pandemic, many seminars, conferences, and talks have been held online. This caters to a wide online audience and is advantageous for easy, rapid, and widespread dissemination of expert opinions and information. The ability to organize large webinars at short notice allows experts to be at the forefront of spreading awareness. In fact, studies have shown that dissemination of scientific journals and articles through social media platforms such as Facebook and Twitter increase the number of downloads and citations [22]. Moreover, guidelines and practice procedures usually used in the hospital or school setting are being shared more during the pandemic, with social media being used as an effective vehicle. Online platforms such as Zoom, Skype, and Whatsapp are complemented by free and simple-to-use collaboration software [23]. Social media has always been known as a form of instant communication and this has been especially highlighted with the onset of the pandemic. More and more of our government agencies, medical societies, and news outlets have embraced this ability to be able to communicate and interact directly with the audience and receive direct feedback. However, a hurdle for the use of social media and as the medium of information dissemination is the possibility that the information may not be current, not subjected to peer review, invalid, not applicable to varying areas, or even incorrect [24]. It is therefore crucial to be able to identify reliable and credible sources of information amongst the abundance of information online.

Findings on the knowledge levels, attitude, and practices may provide valuable insights on how public health initiatives can improve disease prevention during the time of pandemic through strategic behavioral interventions [7]. In terms of knowledge, our study shows that about 78% of medical students had adequate knowledge on COVID-19 infection and prevention. Similar studies also showed good knowledge on COVID-19 infection and prevention among medical students from Pakistan (71.7%) and India (92.7%) [15, 25]. Our results show that in particular, medical students are well-informed regarding the etiology, incubation period, symptoms, preventive measures and complications of COVID-19. However, a relatively lower percentage of respondents were correct regarding the high-risk populations, case fatality rate, and mode of transmission. Local infographics regarding COVID-19 disseminated online usually include the etiology, mode of transmission, incubation period, symptoms, preventive measures, and complications which may explain high knowledge levels for these aspects of COVID-19 [27]. In contrast, high-risk populations and case fatality rate are not usually presented. This may be due to an effort to maintain public alertness in social circles even outside the high-risk group. Furthermore, the COVID-19 case fatality rate may not be usually presented as this statistic varies temporally as the pandemic progresses and spatially among different regions [27]. For mode of transmission, about 23% only considered respiratory droplet as the means for spreading infection. Transmission may also be airborne or through direct contact with infected people and indirect contact with surfaces or objects used by the infected person [28]. This may be due to conflicting information in different sources and lack of conclusive studies. The World Health Organization (WHO) recognizes that airborne transmission of SARS-CoV-2 can occur during medical procedures that generate aerosols [28]. On the other hand, the Centers for Disease Control and Prevention (CDC) states that although there are studies that indicate that inhaling the virus can lead to infection, the relative contribution of this mode of transmission remains unquantified and difficult to establish [29]. Similarly, WHO recognizes direct and indirect contact with infected surfaces as a possible way of spreading the disease whereas CDC emphasizes that current evidence suggests that transmission from contaminated surfaces do not have substantial contribution to new infections [28, 29].

In terms of attitude, our study shows that about 80.68% of medical students have a positive attitude towards COVID-19 infection and prevention. Similar studies also showed a positive attitude towards COVID-19 infection and prevention among medical students from Pakistan (92.5%) and India (80%) [15, 25]. The more positive attitudes among medical students may stem from their consciousness of duty and concern as future physicians [15]. In our study, positive attitude was a reflection of the belief that it was their social responsibility to minimize infection through safety measures, an asymptomatic patient can still transmit the virus to others, hand-washing and wearing of masks are sufficient to prevent infection, the COVID-19 vaccine can aid in protection against infection, the COVID-19 vaccine will be made accessible to most people, and that COVID-19 will be controlled completely. Among these outlooks, it is noteworthy to highlight that almost one-third of the respondents (28.36%) are unsure that the pandemic will be controlled completely. This may be due to the current state of the Philippines and the lack of organizational preparedness to counter the COVID-19 threat. In response to the pandemic, travel restrictions, community quarantine, risk communication, and testing were implemented by the government. However, the slow reinforcement of capacities, especially testing, made it difficult to control the spread of infection in our country [30]. With the unrestrained transmission, this led to longer and stricter lockdowns in the Philippines [31]. Despite this, the Philippines is ranked 25^th^ among countries with the highest number of COVID-19 cases worldwide and 28^th^ among those with the highest number of COVID-19 casualties. Moreover, the country is second in Southeast Asia in both parameters [32].

In terms of practices, our study also shows that about 94.38% of medical students have good practices with regards to COVID-19 prevention. About 96% of the respondents always wear face masks as a precautionary measure. Similarly, 97% of medical students from Afghanistan in another study frequently used face masks [33]. However, for medical students from Jordan in another study, only 9.7% often wore a mask to prevent infection [21]. Differences in local ordinances and culture may account for the stark contrast in the use of face masks. In the Philippines, a directive was released for the strict implementation of mandatory wearing of face masks in public areas in order to prevent the spread of infection [34]. Although not mandatory, the local government also disseminated health advisories on other preventive measures including limiting attendance in huge gatherings, hand-washing or use of alcohol-based sanitizers, avoiding animal contact, observing proper cough etiquette, and avoiding individuals with symptoms of cough and colds [26].

KAP scores were also analyzed with respect to the demographic characteristics to determine possible factors that may influence KAP. Correlational analysis through a Chi-square test only identified sex to be significantly associated with practice wherein females generally had better practice scores than males. This is in congruence with the findings of other studies during the COVID-19 pandemic wherein females were found to be more likely to engage in practices such as social distancing and hand hygiene [15, 35]. Previous studies which were conducted during different disease outbreaks also showed similar results and have attributed their findings to the fact that females are more likely to have better hand hygiene and more likely to wear masks [36, 37]. The binary logistic regression also showed that females are 1.65 times more likely to have adequate knowledge and attitude. This finding is also consistent with other studies that show that women are more likely to perceive COVID-19 as a very serious health problem, agree with restraining public policy measures and comply with them [38, 39]. Women were reported to rely on information from data-driven sources more than men and are more influenced by all four internal sources which are health history, anxiety, feeling responsible for others, and feeling responsible for oneself. The tendency to listen to data-driven sources and tendency to consult internal sources positively correlates with preventive health practices, suggesting women are more likely to listen to sources that motivate compliance with preventive COVID-19 health practices [35].

Another finding from the binary logistic regression was that 4^th^ year students are 1.35 times more likely to have adequate knowledge. Other studies had similar findings wherein academic level was a significant factor associated with the level of knowledge [39, 40, 41]. Academic level and exposure to clinical practice was thought to influence the level of students’ knowledge as fifth-year students gain more experience in clinical training [40]. Second year students were also found to be 1.74 times more likely to have a positive attitude. This is in contrast to the study done by Khasawneh et al. in Jordan wherein they have identified a statistically significant relationship between the academic level and attitude in COVID-19 practices [21].

Being within Cebu City during the quarantine period was found to increase the likelihood of good practice by 1.11 times. This may be attributed to the fact that Cebu City was one of the COVID-19 hotspots in the country, thus emphasizing the greater need to adhere to COVID-19 prevention measures [42]. The longer and stricter quarantine status and strategies implemented by the national and local governments together with the support of the Cebuano community contributed to a significant drop in the number of new COVID-19 cases. The city was able to contain the surge and was on its way to flatten the epidemic curve [42].

Lastly, students whose hometowns are within the city had an increase in the likelihood of positive attitude by 1.21 times. In a study by Prasetyo et al. on the perceived effectiveness of COVID-19 prevention measures among Filipinos during enhanced community quarantine in Luzon, it was found that perceived vulnerability and perceived severity had significant indirect effects on an individual’s attitude towards COVID-19 and intention to follow guidelines. Since Cebu City was one of the COVID-19 hotspots in the country in 2020, Cebuanos may be more likely to be willing to engage and adhere to quarantine protocols to prevent another surge of cases. Cebuanos may also have a more optimistic attitude towards the pandemic since they were able to control the first surge of cases in 2020 and people are more likely to comply with rules and guidelines when they have a positive attitude [43]. Students whose hometown are within the city also are likely to live with older people, such as their parents and other relatives, while those whose hometown are outside Cebu are likely to live separately in dormitories or apartments. Therefore, students who live with someone who is vulnerable to COVID-19 may feel a greater social responsibility to take safety measures in controlling the spread of the virus [44]. However, all confidence interval in the binary logistic regression results crosses 1 (e.g. 95% CI for sex at 0.979, 2.769), which then implies that there is no difference between the groups considered in the study.

The role of knowledge and attitude in influencing practices on COVID-19 prevention was also reaffirmed wherein both knowledge and attitude are correlated with practice. This trend was also seen in other studies [45, 46]. With knowledge having an impact on our compliance to COVID-19 precautionary measures, communication to the public on infection and prevention should be evidence-based and delivered in an understandable language. Proper knowledge would then aid the public to make informed decisions on health issues [47]. Since attitude was also found to play a role in shaping our COVID-19 infection and prevention practices, this would imply that the rationale behind these practices as well as their effectiveness should also be part of health interventions. In order to urge the public to perform these precautionary measures consistently, they should believe that doing so would be of benefit to them [7].

## CONCLUSION

Overall, medical students of CIM have adequate knowledge, positive attitude, and good practices towards COVID-19 infection and prevention measures. Demographic factors may affect KAP wherein females have a significant association with better practices than male counterparts. Knowledge and attitude also play a role in shaping practices which is why health interventions should promote accurate, evidence-based information as well as the effectiveness of precautionary measures that are being implemented.

## RECOMMENDATIONS

For further studies, inclusion of students in other medical schools in the Philippines is recommended to encompass a broader population for a more accurate reflection of KAP among medical students. Another aspect that may be looked into would be extending the scope of the study beyond medical students to the general public, especially residents of highly affected areas of Cebu. A study that would focus on specialized topics on prevention measures such as adherence to quarantine and vaccination may also be beneficial. The results of this study showed that most of the participants’ source of information came from social media. Further dissection of this result would be helpful to determine whether information is reliable or not.

## Data Availability

All data produced in the present work are contained in the manuscript.

## ACKNOWLEDGEMENT

First and foremost, the researchers of this study would like to express their gratitude to the following people who have journeyed with the researchers all throughout.

To their research instructor, Dr. Maria Philina Villamor, for sharing her time, effort, and expertise despite her busy schedule during consultations, imparting her knowledge, recommendations and suggestions to the researchers, and for giving the necessary information which helped the researchers in the study. Throughout the process, she was there to guide and help the researchers.

To their statistician, Mr. Mark Borres, for analyzing the data gathered and for interpreting the results comprehensively which allowed the researchers to reach conclusions about the study.

To the families and friends of the researchers, for their never-ending love and support emotionally, morally, and financially, and their encouragement in any way possible.

And lastly to the Almighty God the Father for his spiritual guidance and bountiful blessings. For giving the researchers the strength to face every battle of the way and to overcome every challenge given throughout the journey.

